# On the association between SARS-COV-2 variants and COVID-19 mortality during the second wave of the pandemic in Europe

**DOI:** 10.1101/2021.03.25.21254289

**Authors:** Katarzyna Jabłońska, Samuel Aballéa, Pascal Auquier, Mondher Toumi

## Abstract

**BACKGROUND:** Preliminary clinical evidence suggests an increased COVID-19 mortality associated with the variant of concern 20I/501Y.V1. The evidence outside the UK and a real-world comparison of variants spread and mortality is sparse. This study aims at investigating the association between COVID-19 mortality and SARS-COV-2 variants spread during the second wave of the COVID-19 pandemic in Europe.

**METHODS:** For 38 European countries, publicly available data were collected on numbers of COVID-19 deaths, SARS-COV-2 variants spread through time using Nextstrain classification and countries’ demographic and health characteristics. The cumulative number of COVID-19 deaths and the height of COVID-19 daily deaths peak during the second wave of the pandemic were considered as outcomes. Pearson correlations and multivariate generalized linear models with selection algorithms were used.

**FINDINGS:** The average proportion of 20I/501Y.V1 variant (B.1.1.7) was found to be a significant predictor of cumulative number of COVID-19 deaths within two months before the deaths peak and between 1 January – 25 February 2021, as well as of the deaths’ peak height when calculating the proportion during the second wave and the pre-peak period. The average proportion of 20A.EU2 variant (S:477N) was a significant predictor of cumulative COVID-19 deaths in the pre-peak period.

**INTERPRETATION:** Our findings suggest that the spread of a new variant of concern 20I/501Y.V1 had a significant impact on the mortality during the second wave of COVID-19 pandemic in Europe and that proportions of 20A.EU2 and 20I/501Y.V1 variants were associated with increased mortality in the initial phase of that wave.

**Research in context:** *Evidence before this study:* Emerging evidence suggests that the new variant of concern 20I/501Y.V1 (B.1.1.7) may be associated with an increased risk of death. The 20A.EU2 variant (S:447N), observed firstly in July 2020 in western Europe, was found to be capable of increasing SARS-COV-2 infectivity. The evidence outside the UK is still sparse, same as a real-world comparison of distinct variants spread and mortality through time.

*Added value of this study:* In this study we investigated whether the change of the proportion of any SARS-COV-2 variant, including 20I/501Y.V1 and 11 other variants identified by Nextstrain up to 25 February 2021, has an association with COVID-19 cumulative mortality or with the height of the second wave COVID-19 mortality peak.

*Implications of all the available evidence:* Our findings shed light on the causes of the increased COVID-19 mortality during the second wave of the pandemic in Europe. It shows the need for early containment strategies when the variant 20I/501Y.V1 emerges. These findings also support the need for systematic SARS-CoV-2 regular genome sequencing to control the COVID-19 pandemic.

## Introduction

After a year since the coronavirus infectious disease 2019 (COVID-19) outbreak has been announced as a pandemic by the World Health Organization (WHO) on 11 March 2020,^1^ there is still a growing interest in monitoring the virus spread and investigating factors which can have an impact on disease mortality.^2-5^ The situation seems to have worsened in countries where new variants emerged (Brazil, UK, US).^6-8^ In the UK the new SARS-CoV-2 “variant of concern” (VOC) has been identified on 20 September 2020 (20I/501Y.V1 or B.1.1.7 mutation or UKV).^7^

As reported by the Public Health England (PHE), an increased risk of hospitalization and transmissibility has been detected for VOC B.1.1.7.^9^ Emerging evidence suggests that the “UKV” may be associated with an increased risk of death.^10-13^ Apart from that, earlier identified variant 20A.EU2 (mutation S:447N), observed firstly in July 2020 in western Europe, was found to be capable of increasing SARS-COV-2 infectivity.^14,15^ Besides newly identified mutations, it cannot be ignored that SARS-COV-2 variants developed during the 1^st^ wave can also have impact on the mortality during the second wave.

Since the preliminary clinical evidence supports the hypothesis of an increased mortality associated with the 20I/501Y.V1 VOC in the UK, a broader view on the problem is of public health interest. The evidence on the actual impact of the UKV outside the UK is still sparse. Therefore, in this study we collected country-level data on the COVID-19 mortality during the second (winter) wave of the pandemic in Europe and investigated whether the change of the proportion of any SARS-COV-2 variant, including the UKV and 11 other variants identified by Nextstrain up to 25 February 2021^16,17^, has an association with COVID-19 cumulative mortality or with the height of the second wave COVID-19 mortality peak.

This study aims at detecting potential association between COVID-19 mortality and proportion of SARS-COV-2 variants through the second wave of the pandemic in Europe with the use of multivariate regression models. To the best of our knowledge, the problem has not been investigated so far in this broader context. This analysis enables us to fill the gap in evidence and shed light on the causes of the increased COVID-19 mortality during the second wave and through the first months of 2021 in Europe.

## Methods

### Data collection

A total of 38 European countries were included in the analysis. The cumulative number of COVID-19 deaths during the second (winter 2020/2021) wave of COVID-19 pandemic was the primary outcome of interest. The secondary outcome of interest was the height of COVID-19 daily deaths peak during the second wave, defined as maximum daily reported number of people who died due to COVID-19 per country, considering the period from start of the second wave to 25 February 2021.

As the start of the second wave is not easily determined, we approximated this date as the median date between 1^st^ wave daily deaths peak height (no later than mid-June 2020) and second wave daily deaths peak height (no sooner than mid-August 2020) when a minimum number of deaths per day ±0.1 deaths per 1 mln inhabitants was observed.

Values of cumulative number of deaths and deaths peak height were divided by the number of inhabitants of a given country and reported as number of deaths per 1 million inhabitants.

The main explanatory variables of interest, assumed to have a potential association with the above outcomes, were average proportions of SARS-COV-2 sequences among identified sequences in the same period used to form Nextstrain clades. Twelve (12) clades (19A, 19B, 20A, 20A.EU1, 20B, 20C, 20D, 20E.EU2, 20G, 20H/501Y.V2, 20I/501Y.V1, 20J/501Y.V3) were identified by Nextstrain from December 2019 to March 2021 in the European region. ^16,17^

Four time periods were considered to analyze the association between outcomes and average proportions of each clade (virus variant) through time:

- From start of the second wave up to second wave COVID-19 daily deaths peak,
- Within two months before reaching the second wave COVID-19 daily deaths peak,
- In the period between 1 January 2021 to 25 February 2021,
- From start of the second wave up to 25 February 2021.

Additional covariates considered in the analysis were: country population size, cumulative number of COVID-19 deaths during the 1^st^ wave of the pandemic, cumulative number of vaccinated people up to the end of considered period, all beds capacity (number of hospital bed units), percentage of population living in metropolitan cities with more than 1 million inhabitants, percentage of population aged 65 or more, prevalence of diabetes, cancer and obesity (2017) and gross domestic product (GDP) per capita (2019). These factors were assessed as significantly impacting the risk of severe illness or mortality from COVID-19 in the literature. ^4,18-20^

Variables indicating deaths, vaccinated people and beds capacity were considered in relation to the population size of a country.

### Data sources

Data on COVID-19 deaths, infections and beds capacity were obtained from Institute for Health Metrics and Evaluation (IHME) on 25 February 2021.^21^ Daily number of deaths were recalculated using 7-day moving average to minimize bias related to possible reporting fluctuations.

A dataset of 3971 SARS-CoV-2 virus strains identified between December 2019 and March 2021, used to form clades of virus variants, was downloaded from the Global Initiative on Sharing All Influenza Data (GISAID) database (https://www.gisaid.org) on 12 March 2021.^22^ In particular, the dataset included information on date and country where a given strain was observed, as well as GISAID and Nextstrain clade to which a given strain was classified. We assumed that if a strain was observed on a given date, it could be observed in a range of ±14 days from the observation date. Since the data are not reported daily, the assumption helps to avoid fluctuations.

Data on number of vaccinated people were taken from Our World In Data.^23^ Data on population size were taken from Worldometer.com.^24^ Data on the population living in metropolitan areas were downloaded from Eurostat and other sources. ^4,19,20,25^ Data on prevalence of diseases, gross domestic product (GDP) and population age were downloaded from Our World In Data and World Bank websites. ^26,27^

### Statistical analysis

Descriptive statistics on outcomes and explanatory variables (N, mean, standard deviation, minimum, maximum) were calculated across European countries for each of considered time periods.

Pearson correlations between average virus variants proportions (raw proportions included in a range [0, 1]) and cumulative deaths and/or deaths peak height were analyzed. Then, multivariate general linear models (GLMs) with a normal distribution function and logistic link function were run using stepwise selection algorithms, to select significant variables and avoid potential bias due to relatively low sample size. A criterion of having p-value lower than 0·1 was applied for each variable to stay in and to enter a model. Variants with mean proportion across countries lower than 0·01 during a considered period were not included in the multivariate analysis. If more than one variant proportion were found significant in a model, Pearson correlations between them were verified.

Additionally, sensitivity analysis of covariates selection was performed using the genetic algorithm applied on the GLM models, with the best model selected based on the value of Akaike’s Information Criterion (AIC) corrected for small sample sizes (AICC). Models with only main effects were considered.

Given a country-level analysis, Moran’s I and Geary’s C statistics^28,29^ were produced to check the spatial autocorrelation in values of outcomes and to determine if it should be considered in final models.

For all analyses, a p-value lower than 0·05 was considered as statistically significant.

Analyses were performed using SAS 9.4 software. R 3.6.2 was used to apply the genetic algorithm.

## Results

### Descriptive statistics

Descriptive statistics of the average proportion of virus variants and outcomes for each of considered time periods are presented in Table 1. Plots presenting the change of proportion of 20I/501Y.V1 variant through time is presented on Figure 1, and on Figure 2 for the UK.

**Table 1.** **Descriptive statistics on variants proportions and outcomes**

|  | N | From the second wave start to the peak |  | During 2 months before the peak |  | From 1 January to 25 February 2021 |  | From the second wave start to 25 February 2021 |  |
| --- | --- | --- | --- | --- | --- | --- | --- | --- | --- |
|  |  | Mean (SD) | Min-Max | Mean (SD) | Min-Max | Mean (SD) | Min-Max | Mean (SD) | Min-Max |
| Proportion of 19A | 38 | 0 (0-01) | 0 - 0-04 | 0 (0) | 0 - 0-03 | 0 (0-01) | 0 - 0-03 | 0 (0-01) | 0 - 0-03 |
| Proportion of 19B | 38 | 0 (0-01) | 0 - 0-03 | 0 (0-01) | 0 - 0-05 | 0 (0) | 0 - 0-01 | 0 (0) | 0 - 0-02 |
| Proportion of 20A | 38 | 0-19 (0-19) | 0 - 0-76 | 0-22 (0-22) | 0 - 0-72 | 0-20 (0-18) | 0 - 0-64 | 0-21 (0-17) | 0 - 0-58 |
| Proportion of 20A.EU2 | 38 | 0-10 (0-16) | 0 - 0-63 | 0-10 (0-18) | 0 - 0-80 | 0-06 (0-11) | 0 - 0-43 | 0-09 (0-14) | 0 - 0-57 |
| Proportion of 20B | 38 | 0-28 (0-24) | 0 - 1 | 0-21 (0-25) | 0 - 1 | 0-12 (0-15) | 0 - 0-59 | 0-24 (0-21) | 0 - 0-88 |
| Proportion of 20C | 38 | 0-01 (0-02) | 0 - 0-07 | 0-01 (0-03) | 0 - 0-20 | 0 (0-01) | 0 - 0-07 | 0-01 (0-01) | 0 - 0-07 |
| Proportion of 20D | 38 | 0-03 (0-08) | 0 - 0-41 | 0-04 (0-12) | 0 - 0-51 | 0-02 (0-11) | 0 - 0-68 | 0-03 (0-09) | 0 - 0-51 |
| Proportion of 20E.EU1 | 38 | 0-18 (0-21) | 0 - 0-71 | 0-22 (0-25) | 0 - 0-87 | 0-15 (0-18) | 0 - 0-55 | 0-17 (0-19) | 0 - 0-66 |
| Proportion of 20G | 38 | 0-01 (0-04) | 0 - 0-27 | 0 (0) | 0 - 0 | 0 (0-01) | 0 - 0-02 | 0-01 (0-04) | 0 - 0-23 |
| Proportion of 20H/501Y.V2 | 38 | 0 (0) | 0 - 0 | 0 (0) | 0 - 0 | 0-01 (0-03) | 0 - 0-16 | 0 (0-01) | 0 - 0-04 |
| Proportion of 20I/501Y.V1 | 38 | 0-03 (0-06) | 0 - 0-28 | 0-07 (0-17) | 0 - 0-64 | 0-3 (0-22) | 0 - 0-91 | 0-09 (0-08) | 0 - 0-39 |
| Proportion of 20J/501Y.V3 | 38 | 0 (0) | 0 - 0 | 0 (0) | 0 - 0 | 0 (0-01) | 0 - 0-05 | 0 (0) | 0 - 0-01 |
| Cumulative number of deaths [per 1 mln inhabitant] | 38 | 431-23 (262-01) | 24-12 - 1022 | 351-89 (209-67) | 24-12 - 815-25 | 327-04 (208-49) | 1-59 - 860-31 | 851-30 (466-45) | 52-59 - 1734 |
| Second wave deaths peak height [per 1 mln inhabitants] | 38 | .. | .. | .. | .. | .. | .. | 11-98 (6-59) | 1-20 - 28-32 |

Abbreviations: SD, standard deviation; Min, minimum; Max, maximum; mln, million.

**Figure 1.**
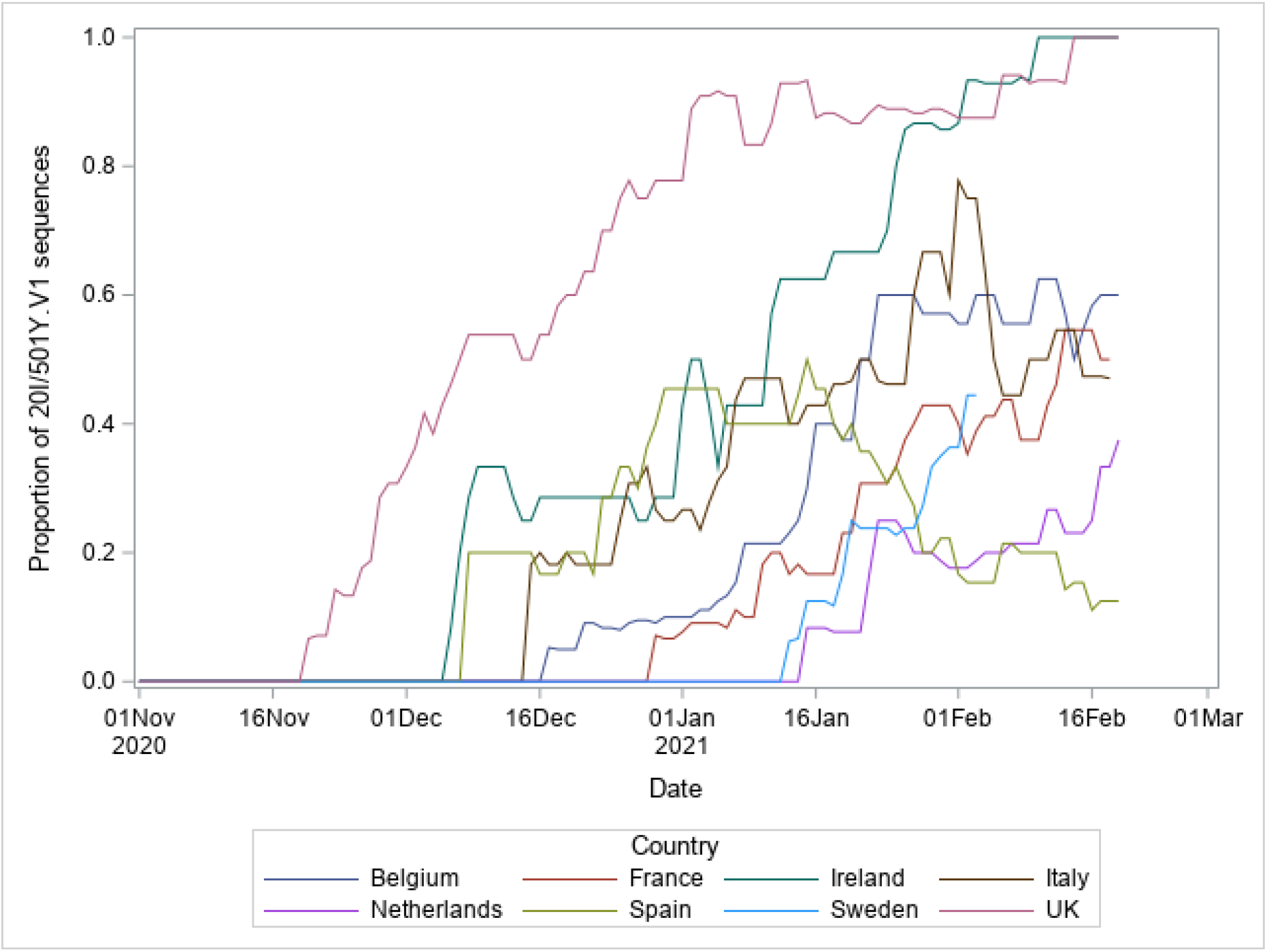
**Proportion of 20I/501Y.V1 sequences among all variants sequences when forming Nextstrain clades (for 8 countries)**

**Figure 2.**
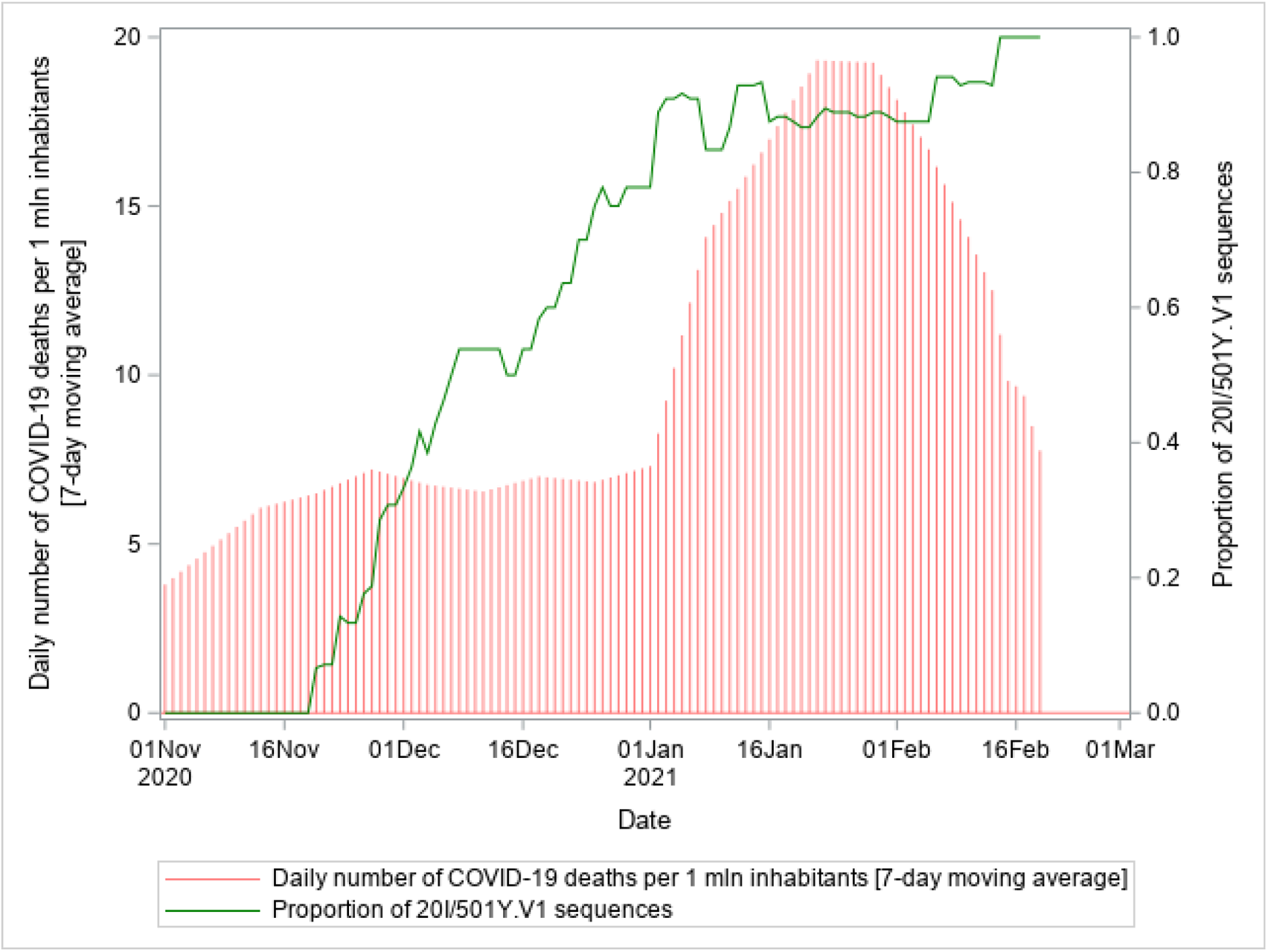
**Proportion of 20I/501Y.V1 variant versus daily number of COVID-19 deaths through time in the UK (1 November – 25 February 2021)**

Moran’s I and Geary’s C statistics for study outcomes indicated a significant spatial autocorrelation in almost all cases (Table 2), therefore all final GLMs selected with the use of selection algorithms were further accounted for the spatial autocorrelation.

**Table 2.** **Spatial autocorrelation across study outcomes**

|  | Time period | Coefficient | Observed | Expected | SD | P value |
| --- | --- | --- | --- | --- | --- | --- |
| Cumulative number of deaths [per 1 mln inhabitant] | From the second wave start to the second wave peak | Moran's I | 0-0298 | -0-027 | 0-0253 | 0.0249 |
|  |  | Geary's c | 0-8201 | 1-000 | 0-0761 | 0.0180 |
|  | During two months before the second wave peak | Moran's I | 0-0459 | -0-027 | 0-0253 | 0.0040 |
|  |  | Geary's c | 0-8949 | 1-000 | 0-0761 | 0.1673 |
|  | Between 1 January - 25 February 2021 | Moran's I | 0-0123 | -0-027 | 0-0253 | 0.1207 |
|  |  | Geary's c | 0-8178 | 1-000 | 0-0761 | 0.0167 |
|  | From the second wave start to 25 February 2021 | Moran's I | 0-0852 | -0-027 | 0-0253 | <0.0001 |
|  |  | Geary's c | 0-8393 | 1-000 | 0-0761 | 0.0347 |
| Second wave deaths peak height [per 1 mln inhabitants] |  | Moran's I | 0.0584 | -0-027 | 0-0253 | 0-0007 |
|  |  | Geary's c | 0.8476 | 1-000 | 0-0761 | 0-0452 |

Abbreviations: SD, standard deviation; mln, million.

### Base case analysis

#### Pearson correlations

Significant positive correlations between the average proportion of 20I/501Y.V1 variant of concern and cumulative number of COVID-19 deaths were observed during the time periods: from start of the second wave to the second wave peak (0·39, p=0·017), within two months before the second wave peak (0·32, p=0·047) and between 1 January – 25 February 2021 (0·47, p=0·003).

A significant negative correlation between the average proportion of 20A variant and second wave deaths peak height was observed for the period from start of the second wave up to the peak (−0·47, p=0·002), whereas its correlation with cumulative number of deaths during that period was close to reaching the significance (−0·32, p=0·051). Considering the entire second wave period, a negative correlation between the average proportion of 20A variant and second wave deaths peak height was closed to reaching the significance (−0·30, p=0·066).

No other significant (p<0·05) or close to significant (p<0·1) correlations were observed for any variants within all considered time periods.

#### Multivariate analysis

##### From second wave start to the deaths peak

The average proportion of 20A.EU2 variant was found to be significant in the GLM model with stepwise selection of cumulative number of deaths during the period from second wave start to the deaths peak, with a positive estimate (1·00, p=0·011; respectively; Table 3). Considering the same period, average proportions of 20I/501Y.V1 and 20A variants were found to be significantly related to the second wave deaths peak height, with a positive estimate for the former and a negative for the latter variant (3·06, p=0·004; -1·00, p=0·035; respectively; Table 3). It should be noted that average proportions of both variants during this period were not correlated (Pearson corr. = -0·03, p=0·85).

**Table 3.** **Results of the GLM model using stepwise covariate selection algorithm for cumulative deaths and second wave deaths peak height, accounting for spatial correlation; from the second wave start to the second wave peak (N=38)**

|  | Estimate | Standard Error | P value |
| --- | --- | --- | --- |
| **Cumulative deaths during the period from the second wave start to the second wave peak** |  |  |  |
| Intercept | 6.3101 | 0.1539 | <0.0001 |
| Average proportion of 20A.EU2 variant | 0.9970 | 0.3703 | 0.0109 |
| GDP per capita [1 mln USD] | -16.8039 | 4.9319 | 0.0017 |
| Cumulative number of vaccinated people before the second wave peak [per 1 mln inhabitants] | 15.1827 | 2.7504 | <0.0001 |
| .. |  |  |  |
| **Second wave deaths peak height** |  |  |  |
| Intercept | 2.0985 | 0.5076 | 0.0002 |
| Average proportion of 20A variant | -0.9974 | 0.4533 | 0.0349 |
| Average proportion of 20I/501Y.V1 variant | 3.0603 | 0.9726 | 0.0035 |
| Percentage of population aged 65 or more | 0.07303 | 0.02495 | 0.0062 |
| Cancer prevalence | -0.4449 | 0.2034 | 0.0359 |

GLM multivariate models with normal distribution and logit link function were used to explore factors associated with COVID-19 cumulative deaths and second wave deaths peak height. The number of cumulative deaths and average variants proportions were calculated from the second wave start to the second wave peak. Each model was run using 38 observations. Models were selected based on the use of stepwise selection algorithm.

##### Within two months before the deaths peak

For the period of two months before the peak, the average proportion of 20I/501Y.V1 and 20A.EU2 variants were selected as significant predictors of cumulative number of deaths during that period (1·41, p<0·001; 0·99, p=0·001; respectively; Table 4), and the proportions were not correlated (Pearson corr. = - 0·18, p=0·28). The 20A.EU2 proportion was also found significant in the model of the deaths’ peak height (1·29, p=0·004; Table 4).

**Table 4.** **Results of the GLM model using stepwise covariate selection algorithm for cumulative deaths and second wave deaths peak height, accounting for spatial correlation; two months before the second wave peak (N=38)**

|  | Estimate | Standard Error | P value |
| --- | --- | --- | --- |
| **Cumulative deaths during two months before the second wave peak** |  |  |  |
| Intercept | 5.9980 | 0.1405 | <.0001 |
| Average proportion of 20A.EU2 variant | 0.9926 | 0.3220 | 0.0041 |
| Average proportion of 20I/501Y.V1 variant | 1.4066 | 0.3311 | 0.0002 |
| GDP per capita [1 mln USD] | -11.9994 | 4.2527 | 0.0079 |
| .. |  |  |  |
| **Second wave deaths peak height** |  |  |  |
| Intercept | 1.8884 | 0.5214 | 0.0009 |
| Average proportion of 20I/501Y.V1 variant | 1.2865 | 0.3726 | 0.0015 |
| Percentage of population aged 65 or more | 0.08907 | 0.02595 | 0.0016 |
| Cancer prevalence | -0.5839 | 0.2038 | 0.0071 |

GLM multivariate models with normal distribution and logit link function were used to explore factors associated with COVID-19 cumulative deaths and second wave deaths peak height. The number of cumulative deaths and average variants proportions were calculated within two months before the second wave peak. Each model was run using 38 observations. Models were selected based on the use of stepwise selection algorithm.

##### 1 January – 25 February 2021

The selected GLM model of cumulative deaths for 1st of January – 25^th^ of February 2021 includes the average proportion of 20I/501Y.V1 variant (1·42, p<0·001; Table 5).

**Table 5.** **Results of the GLM model using stepwise covariate selection algorithm for cumulative deaths between 1 January – 25 February 2021, accounting for spatial correlation (N=38)**

|  | Estimate | Standard Error | P value |
| --- | --- | --- | --- |
| **Cumulative deaths between 1 January – 25 February 2021** |  |  |  |
| Intercept | 4.3707 | 0.5729 | <.0001 |
| Average proportion of 20I/501Y.V1 variant | 1.4179 | 0.3471 | 0.0003 |
| Percentage of population aged 65 or more | 0.06612 | 0.02928 | 0.0304 |
| GDP per capita [1 mln USD] | -11.0302 | 4.5006 | 0.0196 |

GLM multivariate model with normal distribution and logit link function was used to explore factors associated with COVID-19 cumulative deaths. The model was run using 38 observations. The number of cumulative deaths and average variants proportions were calculated from 1 January to 25 February 2021. The model was selected based on the use of stepwise selection algorithm.

##### From second wave start to 25 February 2021

Finally, considering the entire period of the second wave up to 25 February 2021, the average proportion of 20I/501Y.V1 variant was selected as a significant predictor of the deaths peak height (2·37, p=0·023; Table 6), but was not selected into the model of cumulative deaths.

**Table 6.**
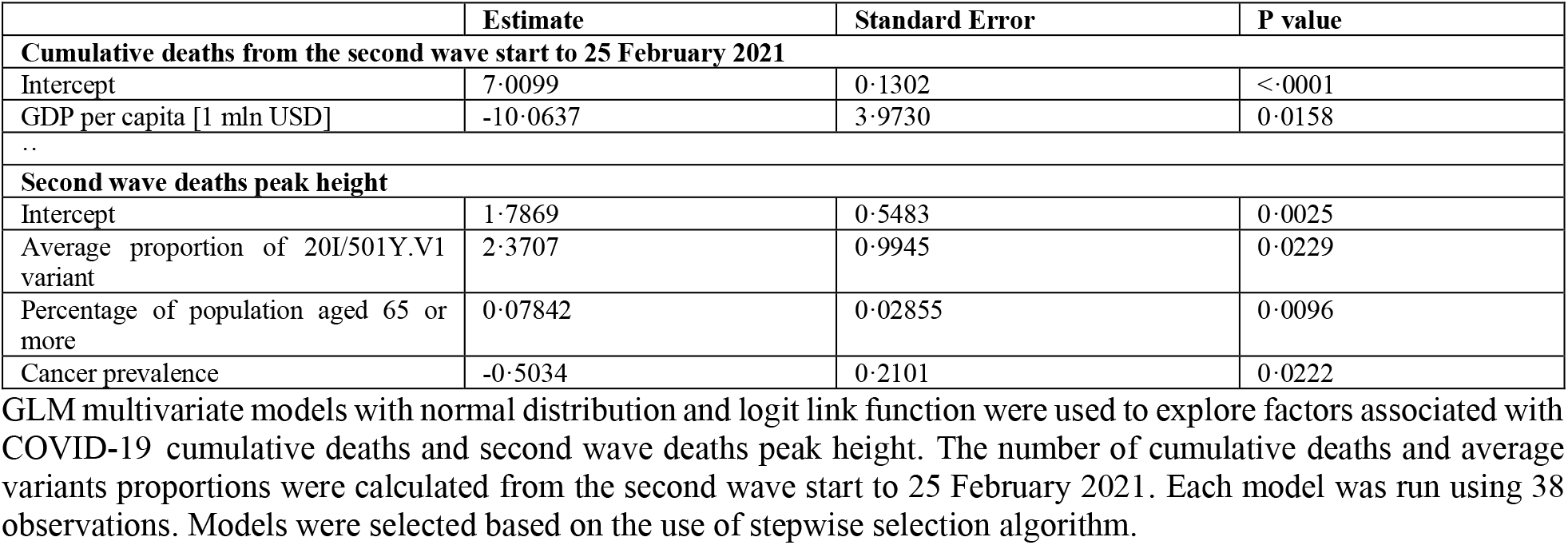
**Results of the GLM model using stepwise covariate selection algorithm for cumulative deaths and second wave deaths peak height, accounting for spatial correlation; from the second wave start to 25 February 2021 (N=38)**

|  | Estimate | Standard Error | P value |
| --- | --- | --- | --- |
| **Cumulative deaths from the second wave start to 25 February 2021** |  |  |  |
| Intercept | 7.0099 | 0.1302 | <.0001 |
| GDP per capita [1 mln USD] | -10.0637 | 3.9730 | 0.0158 |
| .. |  |  |  |
| **Second wave deaths peak height** |  |  |  |
| Intercept | 1.7869 | 0.5483 | 0.0025 |
| Average proportion of 20I/501Y.V1 variant | 2.3707 | 0.9945 | 0.0229 |
| Percentage of population aged 65 or more | 0.07842 | 0.02855 | 0.0096 |
| Cancer prevalence | -0.5034 | 0.2101 | 0.0222 |

GLM multivariate models with normal distribution and logit link function were used to explore factors associated with COVID-19 cumulative deaths and second wave deaths peak height. The number of cumulative deaths and average variants proportions were calculated from the second wave start to 25 February 2021. Each model was run using 38 observations. Models were selected based on the use of stepwise selection algorithm.

### Sensitivity analysis

#### Cumulative deaths

Considering periods from second wave start to the peak, as well as between 1 January – 25 February 2021, same models of cumulative deaths were selected by the genetic algorithm and the stepwise algorithm, suggesting their best fit based on the AICC criterion.

For the period of two months before the deaths peak, genetic algorithm selected a similar model of cumulative deaths as the stepwise algorithm but with one additional variable, percentage of people aged 65 or more. AICC of the model was only slightly better than for the stepwise model (506·70 vs 506·73). However, proportions of 20I/501Y.V1 and 20A.EU2 variants remained significant (1·49, p<0·001; 0·89, p=0·009; respectively; Table 7).

**Table 7.** **Sensitivity analysis results of the GLM model using genetic covariate selection algorithm for cumulative deaths during two months before the second wave peak, accounting for spatial correlation (N=38)**

|  | Estimate | Standard Error | P value |
| --- | --- | --- | --- |
| **Cumulative deaths during two months before the second wave peak** |  |  |  |
| Intercept | 5.1807 | 0.5837 | <.0001 |
| Average proportion of 20A.EU2 variant | 0.8938 | 0.3197 | 0.0086 |
| Average proportion of 20I/501Y.V1 variant | 1.4900 | 0.3368 | <.0001 |
| Percentage of population aged 65 or more | 0.04691 | 0.03145 | 0.1453 |
| GDP per capita [1 mln USD] | -13.9679 | 4.9446 | 0.0080 |

GLM multivariate model with normal distribution and logit link function was used to explore factors associated with COVID-19 cumulative deaths. The model was run using 38 observations. The number of cumulative deaths and average variants proportions were calculated within two months before the second wave peak. The model was selected based on the use of genetic selection algorithm.

For the period from second wave start to 25 February 2021, the genetic algorithm selected a similar model of cumulative deaths as the stepwise algorithm, with GDP per capita (1 mln USD) and additionally, the percentage of people aged 65 or more (Table 8).

**Table 8.**
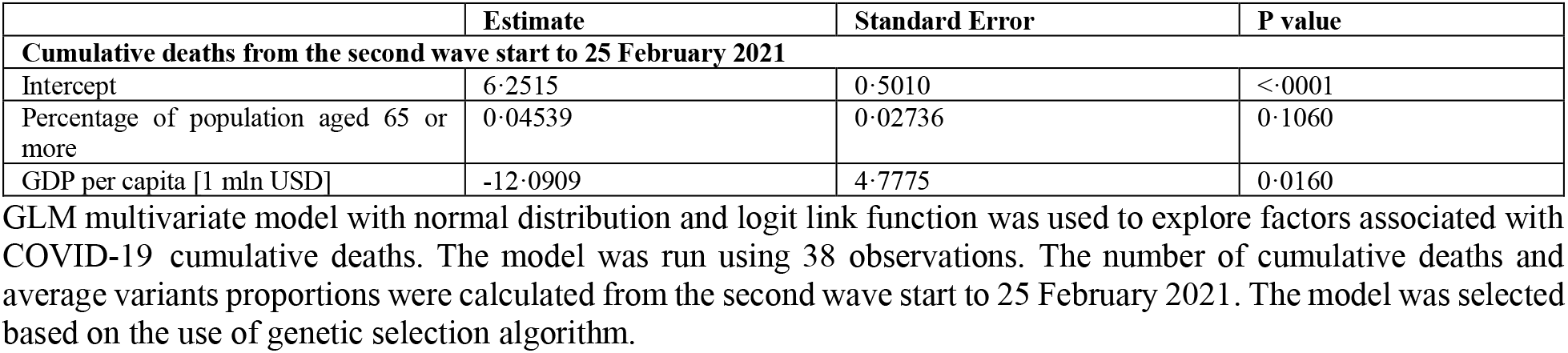
**Sensitivity analysis results of the GLM model using genetic covariate selection algorithm for cumulative deaths from the second wave start to 25 February 2021, accounting for spatial correlation (N=38)**

#### Deaths peak height

For all considered periods, the genetic algorithm selected the same models of the COVID-19 deaths’ peak height as the stepwise algorithm.

## Discussion

In this study we investigated the association between the change of SARS-COV-2 variants proportions through time and COVID-19 cumulative mortality, and the height of the second wave COVID-19 mortality peak. The latter outcome is an indicator of mortality magnitude which can be viewed as less subject to deviations from between-country differences in reporting, not depending on the date up to which the analysis is performed and enabling to assess the overall capacity of healthcare systems.^4^

Our study provides evidence that higher proportion of the VOC 20I/501Y.V1 (B.1.1.7) across countries is associated with higher COVID-19 mortality peak and cumulative mortality during the second wave of the pandemic in Europe. An increase of 0.1 in the proportion of 20I/501Y.V1 variant, considering the pre-peak period, was found to be associated with 35·8% increase in the height of the second wave peak. During the period from 1 January to 25 February 2021, an increase of 0.1 in the proportion of the same VOC was related with a 15·3% increase in the cumulative number of deaths during that period.

These results support previous findings suggesting the increased risk of dying due to B.1.1.7 variant in the UK. The UK’s governmental New and Emerging Respiratory Virus Threats Advisory Group (NERVTAG) reports the results of the matched cohort analysis in which a death risk ratio for VOC-infected individuals compared to non-VOC was 1·65 (95% confidence interval (CI) = [1·21; 2·25]).^13^ Results of a case control study conducted by Challen (et al., 2021) suggest that the mortality hazard ratio associated with infection with VOC B.1.1.7 was 1·64 (95% CI = [1·32; 2·04]) compared to infection with previously circulating variants.^12^ Wallace and Ackland (2021) compared the number of deaths in the UK detected before and after new VOC detection. In early-December 2020 deaths ratio was significantly higher compared to the ratio in October-November 2021, especially in regions affected by the VOC B.1.1.7.^11^

Results of the current study also suggest that the higher proportion of 20A.EU2 variant (mutation S:447N) was associated with increased mortality in the pre-peak phase of the second wave of COVID-19 in the European region. It is complementary to the previous findings that this variant is able to attenuate neutralizing immune responses in humans, studied by Liu (et al., 2020).^14^

The proportion of 20A variant, firstly observed in Europe during the 1^st^ wave of the pandemic (February 2020), was found to be negatively associated with the deaths peak height considering the pre-peak phase of the second wave. While the proportion of 20I/501Y.V1 was positively associated with this outcome, and since no correlation between proportion of 20A and 20I/501Y.V1 was observed in that period, then it can suggest that higher frequency of variants developed during the 1^st^ wave, such as 20A, could potentially be protective during the pre-peak phase of the second wave, due to the fact that societies could have already gained some level of immunity. However, this hypothesis needs further investigations.

We found GISAID to be a reliable source of data on SARS-COV-2 variants spread. The GISAID aims to rapidly share data from influenza viruses and the coronavirus causing COVID-19, including genetic sequence, as well as clinical and epidemiological data associated with human viruses, in order to help researchers understand viruses evolution and spread during epidemics.^30^ SARS-COV-2 variants’ classification used in this study was developed by Nextstrain group. Nextstrain is an open-source project providing real-time analysis of evolving pathogens of the coronavirus and several other human viruses using publicly available data. Nextstrain sheds light on evolution of SARS-CoV-2 viruses from the ongoing COVID-19 pandemic. Based on the phylogenetic analysis, SARS-CoV-2 variants were grouped into clades.^16,17^ More information on clades were obtained from CoVariants, developed by Emma Hodcroft in 2020. The website provides an overview of SARS-CoV-2 variants of the Nextstrain clades, summarizes information on mutations defining each variant, and describes its evolution through the pandemic.^15^

### Limitations

One of the study limitations is that data on limited number of countries were used. Since the course of the pandemic differs sorely between continents, and so do the virus variants spread, we focused on the European region to avoid data inconsistency issues. However, the number of observations is enough to draw conclusions based on multivariate regression models. Stability of results was tested with sensitivity analyses, being highly consistent with base case findings.

The limited number of covariates were included in the multivariate analysis, while other factors could be potentially influential. The selection of covariates was based on the literature search and only factors which previously were found to have a significant association with COVID-19 mortality were included.

Another limitation concerns the reliability of the data. The proportions of variants were calculated across strains obtained from GISAID and sampling of virus strains may not be equal across countries. However, the number of countries with less than 30 observations was only 5 (13·2%), and the average number of sequences per country in the database was 64.

There also exists the variability of data quality between countries, as well as between-country differences on data on daily deaths due to COVID-19 collection methods. In addition, the date of death occurrence and date of reporting can differ.

### Conclusions

Our findings suggest that the development and spread of a new virus variant of concern 20I/501Y.V1 (B.1.1.7) had a significant impact on the mortality during the second wave of COVID-19 pandemic in Europe. It can contribute to explain the persistent high mortality post peak of the second wave and during the first two months of 2021.

Moreover, the frequency of both the novel VOC, as well as of earlier identified 20A.EU2 (S:447N) variant could have a potential influence on excess mortality during the initial phase of the second wave, before reaching the deaths peak.

## Data Availability

This study involved data freely available in the public domain.

http://www.healthdata.org/covid/

https://www.gisaid.org/

## Authors’ contribution

MT conceptualized the study; KJ, SA and PA validated the study concept. KJ and MT collected and verified the data; KJ and SA developed the methodology; KJ analysed the data; KJ, SA and MT interpreted the results; KJ wrote the first draft of the manuscript; SA, PA and MT reviewed and edited the manuscript; KJ, SA, PA and MT revised the manuscript and approved the final version.

## Acknowledgements

None.

## Ethical approval

Not required. This study does not require ethical approval as it was conducted on country-level data and involved information freely available in the public domain.

## Descriptive statistics

## Base case analysis results

## Sensitivity analyses results

